# Evaluation of a commercial Multiplex Real-Time PCR Assay for orthopoxvirus and monkeypox virus detection, and simultaneous subclade Ib identification

**DOI:** 10.1101/2024.11.20.24317626

**Authors:** Tony Wawina-Bokalanga, Michael Haesler, Prince Akil-Bandali, Rilia Ola-Mpumbe, Eddy Kinganda-Lusamaki, Jean-Claude Makangara-Cigolo, Elisabeth Pukuta-Simbu, Fiston Cikaya-Kankolongo, Nelson Mapenzi-Kashali, Ange Ponga-Museme, Tessa de Block, Raphaël Lumembe-Numbi, Adrienne Amuri-Aziza, Gradi Luakanda, Daan Jansen, Koen Vercauteren, Steve Ahuka-Mundeke, Kai Schuchmann, Placide Mbala-Kingebeni, Jean-Jacques Muyembe-Tamfum

**Author notes:** ***Corresponding author:*** Tony Wawina-Bokalanga, MD, PhD.

## Abstract

Several mpox outbreaks, caused by subclades Ia and Ib monkeypox virus (MPXV), are ongoing in the Democratic Republic of the Congo. The World Health Organization declared the ongoing mpox outbreak a Public Health Emergency of International Concern due to the geographic expansion of subclade Ib. We assessed the performance of a novel multiplex real-time PCR assay designed for MPXV detection and simultaneous subclade Ib identification. This assay demonstrates high accuracy and specificity, underscoring its use for the current mpox outbreak.

## Background

Mpox, a zoonotic disease caused by monkeypox virus (MPXV), is endemic to Western and Central African countries, with the highest prevalence in the Democratic Republic of the Congo (DRC) [1].

Monkeypox virus (MPXV) is a large DNA virus that belongs to the genus *Orthopoxvirus* (OPXV) within the family *Poxviridae*, subfamily *Chordopoxvirinae*. The MPXV genome is linear and consists of double-stranded DNA with a length of approximately 197 kb. It encodes approximatively 190 largely non-overlapping open reading frames containing >180 nucleotides. Like all orthopoxviruses, MPXV has a highly conserved central coding region flanked by variable regions containing inverted terminal repeats [2].

Phylogenetically, MPXV is classified into two major clades: Clade I and Clade II. Both clades have been further subdivided into two subclades, with Clade II divided into subclades IIa and IIb, with IIb variants associated with the 2022 global mpox outbreak [3]. Clade I MPXV is further divided into subclades Ia and Ib, with Ib variants first identified in 2023 [4, 5]. Genomic analysis of subclade Ib revealed a ∼ 1 kbp deletion in the OPG032 [4, 5], which resulted in false negative PCR results for subclade Ib when using the Clade I-specific PCR assay originally developed by Li et al [6].

Due to a rapid and geographic expansion of subclade Ib variants within and beyond Africa, including transcontinental importations to Sweden and Thailand, the World Health Organization (WHO) declared mpox a Public Health Emergency of International Concern once again on 14 August, 2024 [7]. In September 2024, we reported for the first time, a co-circulation of both subclades Ia and Ib in Kinshasa, the capital of the DRC [8].

Following the WHO’s call to manufacturers to expand availability of diagnostic assays for specific identification of subclade Ib [9], Gold Standard Diagnostics (GSD; https://clinical.goldstandarddiagnostics.com/) developed a new multiplex real-time PCR assay for this purpose.

In this study, we assessed the clinical performance of this novel multiplex real-time PCR assay using samples from the 2024 mpox outbreak in the DRC, including skin lesion swabs (vesicles and crusts) and blood.

## Methods

### Design of Orthopoxvirus- and MPXV-primers and probes

Primers and probes identical to those designed for the dual-target multiplex real-time PCR assay “Mplex Mpox, Orthopox” (PPOX6201), launched by GSD during the global mpox outbreak in 2022, were used to detect the *F3L* gene of MPXV and *I7L* gene of OPXV. Specific primers for MPXV subclade Ib were designed using CloneManager version 12 (https://www.scied.com/pr_cmpro.htm), based on publicly available subclade Ib sequences from GISAID (www.gisaid.org), by selecting mutations that displayed 100% sequence similarity. MGB-labeled TaqMan probes were designed to specifically discriminate MPXV sequences containing subclade Ib-specific mutations from those of subclade Ia and Clade II.

After the evaluation of three candidate primer- and probe systems using synthetic DNA templates, the mutation P39H in the orthopoxvirus gene 140 (OPG140^P39H^) was selected for the specific detection of subclade Ib variants.

*In-silico* analysis using Geneious Prime version 2023.2.1 demonstrated the binding of primer and probe sequences (OPXV, Generic MPXV and subclade Ib targets) to all available MPXV sequences on GISAID (n=3642, 111 sequences with low genome coverage were excluded).

### Description of the new multiplex real-time PCR Assay

The “Mplex Mpox, Orthopox, Clade 1b” (PPOX6301) real-time PCR assay from GSD contains specific primers and probes labelled with fluorescent reporter and quencher molecules for amplification and simultaneous detection of OPXV, MPXV and subclade Ib MPXV. These primers and probes target a conserved region for orthopoxviruses, a generic target for MPXV (conserved for both clades I and II), as well as a specific target for the subclade Ib variant (Table 1).

**Table 1:** PCR targets and probe specifications for OPXV, MPXV-generic and specific subclade Ib.

| Target | Fluorophore (Quencher) |
| --- | --- |
| I7L (Orthopoxvirus) | HEX<sup>TM</sup> (MGBEQ) |
| F3L (MPXV-generic) | FAM<sup>TM</sup> (BHQ1) |
| OPG140<sup>P39H</sup> (specific subclade Ib MPXV) | Texas Red® (MGBEQ) |
| Human RNase P | ATTO425 (BHQ1) |
| Extraction Control | Cy5<sup>TM</sup> (BHQ2) |

Additionally, a primer and probe set for human RNase P gene serves as an internal positive control to monitor sample quality and detect PCR inhibitors. An additional extraction control reagent can be used to detect extraction failures.

### Real-time PCR evaluation and validation of PCR results through sequencing

Evaluation of the new multiplex real-time PCR assay was performed at the Institut National de Recherche Biomédicale (INRB) located in Kinshasa, DRC. All samples tested in this study were collected in DRC between September and October 2024.

In total, we retrospectively analyzed 43 mpox-positive samples previously assigned as subclade Ib through whole genome sequencing using either Illumina or Oxford Nanopore platform, along with a batch of 67 prospective samples that tested mpox-positive using a routine PCR method based on the Centers for Disease Control & Prevention (CDC) MPXV-generic real-time PCR [10], without known subclade. To provide a broader comparison, we also included 87 samples that tested mpox-negative: 37 samples were tested negative using Xpert® Mpox cartridges (GeneXpert system, Cepheid, USA), and 50 using the CDC MPXV-generic real-time PCR protocol [10].

Viral DNA was extracted using either the DaAnGene extraction kit (https://en.daangene.com/products/spin-column-dna-rna-extraction-kit/) or the QIAamp® DNA Mini Kit (Qiagen, Hilden, Germany), following the manufacturer’s instructions. Real-time PCR assays were performed in duplicate and run on Bio-Rad CFX Opus 96 Real-Time PCR system following these conditions: initial denaturation for 120 sec. at 95 °C; and 40 cycles consisting of 5 sec. at 95 °C and 30 sec. at 60 °C.

## Results

All 43 samples previously tested mpox-positive and assigned to subclade Ib were confirmed as subclade Ib using the GSD PCR assay, demonstrating 100% concordance with sequencing data. The quantification cycle (Cq) values for these samples are shown in Table 2. Among the 67 prospective mpox-positive samples, all samples tested positive for mpox, 8 tested subclade Ib positive, and the remaining 59 were tested subclade Ib negative as detailed in Table 3, with results showing 100% concordance with sequencing results and CDC real-time PCR [10].

**Table 2.** Results of the Mplex Mpox, Orthopox, Clade 1b real-time PCR assay, Democratic Republic of the Congo, September - October 2024, mpox-positive samples assigned as subclade Ib through Whole Genome Sequencing (n=43)

| ID | Mplex Mpox, Orthopox, Clade 1b targets and respective Cq values |  |  |
| --- | --- | --- | --- |
|  | OPXV | MPXV | subclade Ib |
| 1V | 21,40 | 21,14 | 22,23 |
|  | 22,11 | 21,71 | 22,71 |
| 2V | 19,25 | 18,55 | 19,92 |
|  | 18,94 | 18,29 | 19,63 |
| 3C | 20,88 | 20,49 | 21,43 |
|  | 20,74 | 20,30 | 21,19 |
| 4 | 33,14 | 34,57 | 34,27 |
|  | 33,63 | 33,94 | 34,45 |
| 5 | 21,01 | 20,76 | 21,54 |
|  | 20,99 | 20,78 | 21,60 |
| 6 | 22,44 | 22,26 | 23,25 |
|  | 22,91 | 22,77 | 23,68 |
| 7V | 18,39 | 17,87 | 18,98 |
|  | 18,37 | 18,02 | 19,06 |
| 8V | 19,06 | 18,55 | 19,59 |
|  | 19,06 | 18,59 | 19,63 |
| 9V | 17,40 | 16,94 | 18,02 |
|  | 17,35 | 17,01 | 17,99 |
| 10 | 17,69 | 17,23 | 18,31 |
|  | 18,04 | 17,48 | 18,51 |
| 11 | 30,07 | 30,21 | 30,99 |
|  | 30,21 | 30,52 | 31,15 |
| 12 | 21,30 | 21,11 | 21,95 |
|  | 21,96 | 21,72 | 22,52 |
| 13V | 18,84 | 18,60 | 19,47 |
|  | 18,99 | 18,67 | 19,48 |
| 14V | 19,33 | 18,85 | 19,89 |
|  | 19,28 | 18,73 | 19,87 |
| 15V | 18,40 | 18,03 | 19,06 |
|  | 18,45 | 18,04 | 19,06 |
| 16 | 26,29 | 26,40 | 27,20 |
|  | 26,35 | 26,39 | 27,11 |
| 17 | 20,83 | 20,39 | 21,50 |
|  | 21,06 | 20,60 | 21,65 |
| 18 | 25,41 | 25,54 | 26,33 |
|  | 25,51 | 25,66 | 26,37 |
| 19V | 20,61 | 20,27 | 21,28 |
|  | 20,34 | 20,04 | 21,05 |
| 20B | 32,52 | 34,34 | 34,24 |
|  | 33,29 | 35,07 | 34,76 |
| 21V | 21,29 | 20,57 | 21,81 |
|  | 21,27 | 20,54 | 21,82 |
| 22 | 19,72 | 19,30 | 20,27 |
|  | 20,13 | 19,63 | 20,51 |
| 23 | 17,15 | 16,87 | 17,64 |
|  | 17,30 | 17,07 | 17,89 |
| 24 | 27,15 | 27,25 | 28,17 |
|  | 27,15 | 27,28 | 28,02 |
| 25V | 21,09 | 20,81 | 21,61 |
|  | 21,01 | 20,64 | 21,50 |
| 26V | 22,36 | 22,13 | 23,39 |
|  | 22,30 | 22,07 | 23,36 |
| 27V | 17,03 | 16,47 | 17,45 |
|  | 16,75 | 16,24 | 17,19 |
| 28 | 26,34 | 26,41 | 27,33 |
|  | 26,30 | 26,39 | 27,13 |
| 29 | 20,19 | 20,11 | 21,26 |
|  | 20,24 | 20,24 | 21,34 |
| 30V | 23,05 | 22,82 | 23,67 |
|  | 23,01 | 22,77 | 23,72 |
| 31V | 18,68 | 18,24 | 19,28 |
|  | 18,61 | 18,16 | 19,24 |
| 32V | 17,33 | 16,88 | 17,82 |
|  | 17,24 | 16,81 | 17,76 |
| 33 | 22,03 | 21,83 | 22,62 |
|  | 22,18 | 22,01 | 22,86 |
| 34 | 19,15 | 18,83 | 19,79 |
|  | 19,14 | 18,91 | 19,82 |
| 35B | 33,58 | 35,75 | 35,84 |
|  | 32,99 | 36,75 | 35,36 |
| 36V | 19,66 | 19,39 | 20,99 |
|  | 19,37 | 19,18 | 20,61 |
| 37V | 22,31 | 22,07 | 23,11 |
|  | 22,15 | 21,90 | 22,95 |
| 38 | 19,34 | 18,85 | 20,06 |
|  | 19,36 | 18,93 | 20,05 |
| 39 | 23,53 | 23,32 | 24,39 |
|  | 23,41 | 23,29 | 24,31 |
| 40V | 16,17 | 15,88 | 16,72 |
|  | 16,15 | 15,81 | 16,52 |
| 41C | 22,28 | 21,89 | 23,09 |
|  | 22,13 | 21,69 | 22,99 |
| 42 | 25,45 | 25,48 | 26,42 |
|  | 25,54 | 25,58 | 26,22 |
| 43 | 25,15 | 25,27 | 26,17 |
|  | 25,24 | 25,32 | 26,28 |
| NC | ND | ND | ND |
|  | ND | ND | ND |
| PC Ia | 19,38 | 18,96 | ND |
|  | 19,46 | 19,08 | ND |
| PC Ib | 22,04 | 21,67 | 22,68 |
|  | 22,06 | 21,71 | 22,68 |
| PC kit | 28,70 | 28,79 | 29,58 |
|  | 28,25 | 28,37 | 29,92 |

V: Vesicle; C: Crust; B: Blood; NC: Negative Control (Water); PC Ia: Positive Control for subclade Ia; PC Ib: Positive Control for subclade Ib; PC kit: Positive Control from the Mplex Mpox, Orthopox, Clade 1b RT-PCR assay. ND: not detected. OPXV: Orthopoxvirus, MPXV: monkeypox virus.

**Table 3.** Results of the Mplex Mpox, Orthopox, Clade 1b real-time PCR, CDC MPXV-generic real-time PCR assay, and subclade assignment of sequencing data, Democratic Republic of the Congo, September - October 2024, prospective samples tested mpox positive (n=67)

| ID | Mplex Mpox, Orthopox, Clade 1b targets and Cq values |  |  | Interpretation | CDC real-time PCR assay | WGS |
| --- | --- | --- | --- | --- | --- | --- |
|  | OPXV | MPXV | subclade Ib |  | MPXV <sup>a</sup> | subclade |
| 1V | 20,43 | 20,10 | ND | Ia or II | 19,53 | Ia |
|  | 20,74 | 20,29 | ND |  | 19,83 |  |
| 2V | 19,11 | 18,65 | ND | Ia or II | 18,50 | Ia |
|  | 19,12 | 18,52 | ND |  | 18,36 |  |
| 3V | 20,02 | 19,41 | ND | Ia or II | 18,88 | Ia |
|  | 19,54 | 19,09 | ND |  | 18,68 |  |
| 4V | 17,82 | 17,31 | ND | Ia or II | 16,74 | Ia |
|  | 17,85 | 17,26 | ND |  | 16,90 |  |
| 5V | 29,89 | 29,95 | ND | Ia or II | 29,39 | Ia |
|  | 29,93 | 29,97 | ND |  | 29,48 |  |
| 6V | 21,66 | 21,22 | ND | Ia or II | 21,51 | Ia |
|  | 21,87 | 21,41 | ND |  | 21,57 |  |
| 7V | 26,48 | 26,56 | 27,39 | Ib | 26,50 | Ib |
|  | 26,59 | 26,67 | 27,37 |  | 26,49 |  |
| 8V | 26,41 | 26,49 | ND | Ia or II | 25,93 | Ia |
|  | 26,27 | 26,40 | ND |  | 25,84 |  |
| 9V | 15,92 | 15,27 | ND | Ia or II | 15,23 | Ia |
|  | 16,06 | 15,45 | ND |  | 15,30 |  |
| 10V | 18,15 | 17,54 | ND | Ia or II | 17,41 | Ia |
|  | 18,21 | 17,66 | ND |  | 17,42 |  |
| 11V | 21,00 | 20,53 | ND | Ia or II | 20,09 | Ia |
|  | 20,96 | 20,45 | ND |  | 20,09 |  |
| 12V | 20,01 | 19,32 | ND | Ia or II | 18,62 | Ia |
|  | 19,92 | 19,35 | ND |  | 18,64 |  |
| 13V | 19,28 | 18,85 | ND | Ia or II | 18,00 | Ia |
|  | 19,09 | 18,62 | ND |  | 17,99 |  |
| 14V | 21,10 | 20,79 | 21,59 | Ib | 20,46 | Ib |
|  | 21,33 | 21,13 | 21,67 |  | 20,54 |  |
| 15V | 18,82 | 18,36 | ND | Ia or II | 17,80 | Ia |
|  | 18,61 | 18,23 | ND |  | 17,77 |  |
| 16V | 30,70 | 31,13 | 31,93 | Ib | 30,27 | Ib |
|  | 31,00 | 30,95 | 32,04 |  | 30,27 |  |
| 17V | 32,79 | 34,30 | ND | Ia or II | 32,33 | Ia |
|  | 32,43 | 35,32 | ND |  | 32,22 |  |
| 18V | 17,88 | 17,13 | ND | Ia or II | 17,06 | Ia |
|  | 17,70 | 17,07 | ND |  | 16,98 |  |
| 19V | 19,07 | 18,45 | ND | Ia or II | 17,79 | Ia |
|  | 19,05 | 18,42 | ND |  | 17,87 |  |
| 20V | 22,52 | 22,23 | ND | Ia or II | 21,76 | Ia |
|  | 22,42 | 22,08 | ND |  | 21,60 |  |
| 21V | 33,30 | 36,14 | ND | Ia or II | 33,12 | Ia |
|  | 33,16 | 37,09 | ND |  | 32,77 |  |
| 22V | 25,38 | 25,00 | ND | Ia or II | 25,06 | Ia |
|  | 25,40 | 25,05 | ND |  | 25,06 |  |
| 23V | 18,35 | 17,97 | ND | Ia or II | 17,52 | Ia |
|  | 18,34 | 17,82 | ND |  | 17,42 |  |
| 24V | 33,27 | 35,08 | 35,55 | Ib | 32,54 | Ib |
|  | 32,79 | 35,29 | 35,36 |  | 31,81 |  |
| 25V | 18,17 | 17,63 | ND | Ia or II | 17,22 | Ia |
|  | 18,47 | 18,52 | ND |  | 17,40 |  |
| 26V | 19,31 | 18,94 | ND | Ia or II | 18,91 | Ia |
|  | 19,55 | 19,02 | ND |  | 18,97 |  |
| 27V | 18,37 | 17,94 | ND | Ia or II | 17,31 | Ia |
|  | 18,18 | 17,72 | ND |  | 17,23 |  |
| 28V | 20,26 | 19,78 | 20,92 | Ib | 19,54 | Ib |
|  | 20,11 | 19,52 | 20,70 |  | 19,41 |  |
| 29V | 18,35 | 17,91 | ND | Ia or II | 17,33 | Ia |
|  | 18,29 | 17,84 | ND |  | 17,29 |  |
| 30V | 20,07 | 19,57 | ND | Ia or II | 19,36 | Ia |
|  | 20,09 | 19,43 | ND |  | 19,40 |  |
| 31V | 17,87 | 17,34 | ND | Ia or II | 16,89 | Ia |
|  | 17,87 | 17,29 | ND |  | 16,77 |  |
| 32V | 18,09 | 17,69 | ND | Ia or II | 17,38 | Ia |
|  | 18,09 | 17,65 | ND |  | 17,32 |  |
| 33V | 24,65 | 24,46 | ND | Ia or II | 24,31 | Ia |
|  | 24,74 | 24,49 | ND |  | 24,31 |  |
| 34V | 28,30 | 28,29 | ND | Ia or II | 27,42 | Ia |
|  | 28,29 | 28,46 | ND |  | 27,64 |  |
| 35V | 32,77 | 33,89 | ND | Ia or II | 32,23 | Ia |
|  | 33,14 | 34,85 | ND |  | 32,07 |  |
| 36C | 22,32 | 22,20 | ND | Ia or II | 21,82 | Ia |
|  | 22,36 | 22,16 | ND |  | 21,62 |  |
| 37V | 17,63 | 17,17 | ND | Ia or II | 16,07 | Ia |
|  | 17,65 | 17,26 | ND |  | 16,22 |  |
| 38V | 16,21 | 15,91 | ND | Ia or II | 14,51 | Ia |
|  | 16,52 | 16,22 | ND |  | 14,97 |  |
| 39V | 17,96 | 17,50 | ND | Ia or II | 16,61 | Ia |
|  | 17,96 | 17,54 | ND |  | 16,56 |  |
| 40B | 31,24 | 31,56 | ND | Ia or II | 30,81 | Ia |
|  | 31,32 | 31,98 | ND |  | 30,94 |  |
| 41V | 23,34 | 23,27 | 24,25 | Ib | 22,66 | Ib |
|  | 23,34 | 23,27 | 24,23 |  | 22,55 |  |
| 42V | 20,67 | 20,37 | ND | Ia or II | 19,44 | Ia |
|  | 20,70 | 20,39 | ND |  | 19,62 |  |
| 43V | 22,59 | 22,62 | ND | Ia or II | 21,37 | Ia |
|  | 22,68 | 22,72 | ND |  | 21,33 |  |
| 44V | 21,03 | 20,79 | ND | Ia or II | 20,12 | Ia |
|  | 21,04 | 20,81 | ND |  | 20,15 |  |
| 45B | 30,68 | 30,77 | ND | Ia or II | 29,94 | Ia |
|  | 31,45 | 31,19 | ND |  | 29,88 |  |
| 46V | 29,17 | 29,37 | ND | Ia or II | 29,03 | Ia |
|  | 29,46 | 29,77 | ND |  | 29,08 |  |
| 47V | 22,87 | 22,86 | ND | Ia or II | 22,08 | Ia |
|  | 22,58 | 22,75 | ND |  | 22,16 |  |
| 48V | 24,00 | 24,12 | ND | Ia or II | 23,39 | Ia |
|  | 23,95 | 24,08 | ND |  | 23,41 |  |
| 49C | 19,22 | 18,75 | ND | Ia or II | 17,90 | Ia |
|  | 19,18 | 18,60 | ND |  | 17,91 |  |
| 50V | 29,24 | 29,32 | ND | Ia or II | 28,57 | Ia |
|  | 29,38 | 29,36 | ND |  | 28,55 |  |
| 51V | 17,99 | 17,56 | ND | Ia or II | 16,41 | Ia |
|  | 18,08 | 17,66 | ND |  | 16,51 |  |
| 52V | 18,41 | 18,10 | ND | Ia or II | 17,52 | Ia |
|  | 18,29 | 17,98 | ND |  | 17,49 |  |
| 53C | 15,87 | 15,42 | 16,34 | Ib | 15,06 | Ib |
|  | 16,13 | 15,81 | 16,57 |  | 15,07 |  |
| 54V | 25,90 | 25,42 | 26,54 | Ia or II | 24,87 | Ia |
|  | 26,00 | 25,33 | 26,31 |  | 24,68 |  |
| 55B | 31,40 | 31,21 | ND | Ia or II | 30,08 | Ia |
|  | 31,23 | 31,29 | ND |  | 30,21 |  |
| 56V | 15,71 | 15,26 | ND | Ia or II | 14,73 | Ia |
|  | 15,64 | 15,16 | ND |  | 14,60 |  |
| 57C | 16,38 | 16,10 | 16,93 | Ib | 15,01 | Ib |
|  | 16,35 | 16,07 | 16,87 |  | 14,92 |  |
| 58V | 18,03 | 17,41 | ND | Ia or II | 16,52 | Ia |
|  | 17,96 | 17,36 | ND |  | 16,44 |  |
| 59C | 18,65 | 18,28 | ND | Ia or II | 17,54 | Ia |
|  | 18,99 | 18,55 | ND |  | 17,69 |  |
| 60V | 28,94 | 29,20 | ND | Ia or II | 28,22 | Ia |
|  | 28,88 | 28,99 | ND |  | b |  |
| 61C | 17,14 | 16,69 | ND | Ia or II | 16,08 | Ia |
|  | 17,02 | 16,52 | ND |  | 16,01 |  |
| 62B | 29,35 | 29,24 | ND | Ia or II | 28,36 | Ia |
|  | 28,99 | 29,01 | ND |  | 28,30 |  |
| 63V | 20,06 | 19,61 | ND | Ia or II | 18,61 | Ia |
|  | 20,01 | 19,57 | ND |  | 18,54 |  |
| 64V | 29,82 | 29,68 | ND | Ia or II | 28,79 | Ia |
|  | 29,62 | 29,44 | ND |  | 29,02 |  |
| 65C | 17,80 | 17,59 | ND | Ia or II | 16,49 | Ia |
|  | 17,70 | 17,35 | ND |  | 16,53 |  |
| 66V | 23,08 | 23,08 | ND | Ia or II | 22,51 | Ia |
|  | 23,03 | 23,01 | ND |  | 22,47 |  |
| 67V | 27,68 | 27,59 | ND | Ia or II | 26,56 | Ia |
|  | 27,55 | 27,47 | ND |  | 27,02 |  |
| NC | ND | ND | ND | Negative | ND | N/A |
|  | ND | ND | ND |  | ND |  |
| PC Ia | 16,69 | 16,23 | ND | Ia or II | 15,93 | Ia |
|  | 16,77 | 16,28 | ND |  | 16,09 |  |
| PC Ib | 18,77 | 18,33 | 19,33 | Ib | 17,52 | Ib |
|  | 18,25 | 17,89 | 18,88 |  | 17,36 |  |
| PC kit | 31,13 | 31,41 | 32,20 | All targets<br>detected | N/A |  |
|  | 31,67 | 32,25 | 32,85 |  |  |  |

V: Vesicle; C: Crust; B: Blood; NC: Negative Control (Water); PC Ia: Positive Control for subclade Ia; PC Ib: Positive Control for subclade Ib; PC kit: Positive Control from the Mplex Mpox, Orthopox, Clade 1b RT-PCR assay. ND: not detected. WGS: Whole Genome Sequencing, OPXV: Orthopoxvirus, MPXV: monkeypox virus. N/A: Not Applicable. CDC: Centers for Disease Control & Prevention, <sup>a</sup> CDC MPXV-generic real time PCR protocol. <sup>b</sup> Insufficient input DNA material.

Among 87 samples initially tested negative for mpox, five tested positive using the new real-time PCR assay, while the remaining 82 samples remained negative. Importantly, these five samples, tested positive with the new real-time PCR assay, had previously tested negative in a subset of 37 samples analyzed using Xpert® Mpox cartridges (GeneXpert system, Cepheid, USA). To address this discrepancy, we performed additional testing on these five samples using the OPXV detection real-time PCR assay [11] and the CDC real-time PCR [10]. Interestingly, the latter also detected MPXV with Cq values similar to those obtained with the GSD PCR assay (Table 4).

**Table 4.** Results of the Mplex Mpox, Orthopox, Clade 1b real-time PCR, and OPXV and MPXV-generic PCR assay, Democratic Republic of the Congo, September - October 2024, false negative samples using Xpert® Mpox cartridges (n=5)

| ID | Mplex Mpox, Orthopox, Clade 1b targets and respective Cq |  |  | Interpretation | RT- PCR assay targets and respective Cq |  |
| --- | --- | --- | --- | --- | --- | --- |
|  | OPXV | MPXV | subclade Ib |  | OPXV <sup>b</sup> | MPXV <sup>a</sup> |
| 1S-V | 18,25 | 17,87 | ND | Ia or II | 18,14 | 17,12 |
|  | 18,44 | 18,15 | ND |  |  |  |
| 2S-V | 14,21 | 15,33 | 16,73 | Ib | 14,67 | 13,24 |
|  | 14,83 | 14,31 | 15,62 |  |  |  |
| 3S-V | 31,73 | 32,23 | 32,53 | Ib | 32,44 | 30,46 |
|  | 31,50 | 32,72 | 32,74 |  |  |  |
| 4S-V | 33,38 | 36,03 | ND | Ia or II | 34,70 | 33,07 |
|  | 34,19 | 37,12 | ND |  |  |  |
| 5S-V | 35,02 | 36,48 | ND | Ia or II | 33,62 | 31,89 |
|  | 35,44 | 37,43 | ND |  |  |  |
| NC | ND | ND | ND | Negative | ND | ND |
|  | ND | ND | ND |  |  |  |
| PC Ia | 17,07 | 16,90 | ND | Ia or II | 16,09 | 15,93 |
|  | 16,52 | 16,32 | ND |  |  |  |
| PC Ib | 22,51 | 22,32 | 23,94 | Ib | 22,21 | 21,08 |
|  | 22,55 | 22,33 | 23,81 |  |  |  |
| PC kit | 33,15 | 32,34 | 33,56 | All targets detected | N/A |  |
|  | 33,07 | 32,33 | 32,85 |  |  |  |

V: Vesicle; NC: Negative Control (Water); PC Ia: Positive Control for subclade Ia; PC Ib: Positive Control for subclade Ib; PC kit: Positive Control from the Mplex Mpox, Orthopox, Clade 1b RT-PCR kit. ND: not detected. OPXV: Orthopoxvirus, MPXV: monkeypox virus. N/A: Not Applicable. <sup>a</sup> CDC MPXV-generic real time PCR protocol ; <sup>b</sup> Real-time PCR assay for OPXV detection, developed by Schroeder et al, 2010.

## Discussion

Here, we assessed the clinical performance of a novel multiplex real-time PCR assay developed for MPXV detection and simultaneous subclade Ib identification. The evaluated real-time PCR assay resulted in 100% concordance (110/110) with sequencing data for patient specimens positive for both subclades Ia and Ib, and 94.2% concordance (82/87) for negative specimens. The positive results obtained with the new assay suggest an enhanced sensitivity in detecting low viral load samples that have been tested false negative using Xpert® Mpox cartridges (GeneXpert system, Cepheid, USA).

The samples used in this study were obtained from patients of various age groups, including both males and females, and were mainly vesicle swabs, with some crust and blood samples. The Cq values measured by the evaluated GSD assay were comparable to those obtained with the CDC MPXV-generic real-time PCR assay and GeneXpert system. Importantly, the GSD assay additionally provides “subclade Ib detection” within the same PCR reaction, even in samples with low viral load, which are often excluded for sequencing.

A limitation of the GSD assay is that it cannot distinguish between MPXV subclade Ia and Clade II. An additional primer probe set would be required, but the number of detection channels of standard real-time PCR cyclers is commonly limited to five channels. Additionally, the WHO specifies that two internal process controls shall be included, leaving only three channels flexibility for detection of OPXV and MPXV target sequences. This limitation is generally acceptable, as subclade Ib is of primary interest in the DRC, and Clade II mpox cases are historically not reported.

## Conclusion

In summary, a total of 197 patient specimens collected during the ongoing mpox outbreak in DRC were included. Among these, 43 mpox-positive samples previously assigned as subclade Ib were all (100%) confirmed as subclade Ib using the new real-time PCR assay. Furthermore, we tested 87 samples that were previously tested negative for mpox, however, five of these yielded positive results with the new assay. We validated these results by retesting the five samples with OPXV-specific and MPXV-generic primers/probes. The GSD real-time PCR assay proved to be highly sensitive and specific, making it suitable for use in the current mpox outbreak.

## Data Availability

All data produced in the present work are contained in the manuscript.

## Ethical statement

Samples were collected as part of a routine countrywide mpox surveillance program and were exempt from informed consent procedures. The study was conducted in accordance with the Declaration of Helsinki, and all samples were anonymized to ensure the highest standards of patient confidentiality. Permission to use anonymized data from the mpox national surveillance activities for this report was granted by the Ethics Committee of the Kinshasa School of Public Health (ESP-UNIKIN, Number ESP/CE/05/2023).

## Acknowledgment

The authors are grateful to Princesse Paku-Tshambu, Sifa Kavira, Jocelyne Tshimbolela-Mianda and Chloé Muswamba.

